# Is there a serum proteome signature to predict mortality in severe COVID-19 patients?

**DOI:** 10.1101/2021.03.13.21253510

**Authors:** Franziska Völlmy, Henk van den Toorn, Riccardo Zenezini Chiozzi, Ottavio Zucchetti, Alberto Papi, Carlo Alberto Volta, Luisa Marracino, Francesco Vieceli Dalla Sega, Francesca Fortini, Gianluca Campo, Marco Contoli, Savino Spadaro, Paola Rizzo, Albert J.R. Heck

## Abstract

Here we recorded serum proteome profiles of 33 COVID-19 patients admitted to respiratory and intensive care units because of respiratory failure. We received, for most patients, blood samples just after admission and at two more later timepoints. We focused on serum proteins different in abundance between the group of survivors and non-survivors and observed that a rather small panel of about a dozen proteins were significantly different in abundance between these two groups. The four structurally and functionally related type-3 cystatins AHSG, FETUB, HRG and KNG1 were all more abundant in the survivors. The family of inter-α-trypsin inhibitors, ITIH1, ITIH2, ITIH3 and ITIH4, were all found to be differentially abundant in between survivors and non-survivors, whereby ITIH1 and ITIH2 were more abundant in the survivor group and ITIH3 and ITIH4 more abundant in the non-survivors. ITIH1/ITIH2 and ITIH3/ITIH4 also did show opposite trends in protein abundance during disease progression. This panel of eight proteins, complemented with a few more, may represent a panel for mortality risk assessment and eventually even for treatment, by administration of exogenous proteins possibly aiding survival. Such administration is not unprecedented, as administration of exogenous inter-α-trypsin inhibitors is already used in the treatment of patients with severe sepsis and Kawasaki disease. The mortality risk panel defined here is in excellent agreement with findings in two recent COVID-19 serum proteomics studies on independent cohorts, supporting our findings. This panel may not be unique for COVID-19, as some of the proteins here annotated as mortality risk factors have previously been annotated as mortality markers in aging and in other diseases caused by different pathogens, including bacteria.

## Introduction

The coronavirus disease 2019 (COVID-19) pandemic caused by severe acute respiratory syndrome coronavirus 2 (SARS-CoV-2) has affected many people with a worrying fatality rate up to 60% for critical cases. Not all people infected by the virus are affected equally. Several parameters have been defined that may influence and/or predict disease severity and mortality, with age, gender, body mass and underlying comorbidities being some of the most well-established. To delineate best treatments and recognize disease severity early on, it would be very helpful to discover markers helping to define disease severity, have prognostic value, and/or predict a specific phase of the disease. Unfortunately, not many prognostic biomarkers are yet available that can distinguish patients requiring immediate medical attention and estimate their associated mortality rates.

Here, we attempted to contribute to this urgent need aiming to find serum biomarkers that may be used to predict mortality in a group of COVID-19 patients. For the present purpose, we prospectively assessed serum protein levels at different time-points by using mass-spectrometry based serum proteomics in a cohort of moderate-to-severe COVID19 patients admitted to hospital because of respiratory failure (ATTAC-Co study – registered at www.clinicaltrial.gov number NCT04343053).

Given the central role of proteins in biological processes as a whole, and in particular in disease, we applied mass spectrometry-based proteomics to identify protein biomarkers that could discriminate between the COVID-19 patients that recovered and those that did not survive. Several proteomic studies have to date investigated the serum or plasma of COVID-19 patients for the most part comparing a cohort of COVID-19 patients to control subjects (no disease) [1, 2], and in particular an extensive study by Demichev et al. [3] has already investigated the temporal aspect of COVID-19 progression in individuals in order to predict outcome and future disease progression. Although their cohort does comprise some patients that did not survive, for the most part the subjects recovered and were discharged. The unique cohort described in our study allows us to focus on survivors compared to non-survivors and to define clinical classifiers predicting outcome by using subjects recovered from the disease as control group.

We did choose a robust data-independent acquisition (DIA) approach to profile the serum of this patient cohort, as this method circumvents the semi-stochastic sampling bias specific to standard shotgun proteomics, and benefits from high reproducibility. The DIA approach, although not novel [4, 5], has recently increased in popularity in part due to new hardware and software solutions [6], but also due to the efforts of the proteomics community to develop DIA setups that do not require a reference spectral library. In this work we chose to exploit the DIA-NN software suite [7], which makes use of deep neural networks and signal correction to process the complex spectral maps that arise from DIA experiments. This results in a reduction of interfering spectra and in confident statistically significant identifications thanks to the use of neural networks to distinguish between target and decoy precursors.

Using this approach, we observed that, strikingly, the group of survivors and non-survivors could be well separated by just a small group of around a dozen highly abundant serum proteins, already at the first timepoints, i.e., shortly after admission to the ICU. This panel of proteins includes mostly functionally related proteins, including all major type 3 cystatins (HRG, FETUB, ASHG1 and KNG1 and several protease inhibitors (SERPINA2, ITIH1, ITIH2), being more abundant in survivors. As this panel is already able to distinguish the patient groups at the early onset it may have good predictive value. Statistically most significant, the type-3 cystatin histidine rich glycoprotein (HRG) and fetuin-B (FETUB) were consistently more abundant in survivors than in non-survivors. These serum proteins have previously been identified as predictors of mortality in patients affected by *S Aureus* bacteremia [8], but also as general mortality markers in studies looking at aging [9]. Therefore, the panel we observe here may not be specific for patients suffering from COVID-19, and be more generally applicable to predict mortality risk [10].

## Methods

### Individual Serum Sample Collection and Chemicals

The present analysis included patients from the “Pro-thrombotic status in patients with SARS-Cov-2 infection” (ATTAC-Co) study [11, 12]. The ATTAC-Co study is an investigator-initiated, prospective, single-centre study recruiting consecutive patients admitted to hospital (University Hospital of Ferrara, Italy) because of respiratory failure due to COVID-19 between April and May 2020. Inclusion criteria were: i) age >18 years; ii) confirmed SARS-CoV-2 infection; iii) hospitalization for respiratory failure; iv) need for invasive or non-invasive mechanical ventilation or only oxygen support. Exclusion criteria were: prior administration of P2Y12 inhibitor (clopidogrel, ticlopidine, prasugrel, ticagrelor) or anticoagulant drugs (warfarin or novel oral anticoagulants), known disorder of coagulation or platelet function and/or chronic inflammatory disease. SARS-CoV-2 infection was confirmed by RT-PCR (reverse transcriptase polymerase chain reaction) assay (Liaison MDX, Diasorin, Saluggia, Italy) from nasopharyngeal swab specimen. Respiratory failure was defined as a PaO2/FiO2 (P/F) ratio ≤200 mmHg. Clinical management was in accordance with current guidelines and specific recommendations for COVID-19 pandemic by Health Authorities and Scientific Societies. Three different blood samplings (at inclusion (t1), after 7±2 days (t2) and 14±2 days (t3)) were withdrawn. Study blood samplings were performed from an antecubital vein using a 21-gauge needle or from central venous line. All patients underwent blood sampling in the early morning, at least 12 hours after last administration of anticoagulant drugs. The first 2 to 4 mL of blood was discarded, and the remaining blood was collected in tubes for serum/plasma collection. The serum and plasma samples were stored at −80 °C. The planned blood sample withdrawals were not performed in case of patient’s death or hospital discharge. The ATTAC-Co study population consists of 54 moderate-to-severe COVID19 patients [11, 12]. The subgroup of interest for the present analysis is selected starting from the 16 cases who died. From the remaining 38 survivors, we identified the 17 cases who best matched in terms of age, clinical history and clinical presentation. This selection was done with the aim to maximize the possibility to identify differences between deceased and survivors and minimizing potential confounding factors. The protocol was approved by the corresponding Ethics Committee (Comitato Etico di Area Vasta Emilia Centro, Bologna, Italy). All patients gave their written informed consent. In case of unconsciousness, the informed consent was signed by the next of kin or legal authorized representative. The study is registered at www.clinicaltrials.gov with the identifier NCT04343053.

### Serum sample preparation

A detergent-based buffer (1% SDC, 10mM TCEP, 10mM Tris, 40mM chloroacetamide) with Complete mini EDTA-free protease inhibitor cocktail (Roche) was added to serum at a 25:1 ratio to enhance protein denaturation and boiled for 5min at 95C. 50mM ammonium bicarbonate was added and digestion was allowed to proceed overnight at 37°C using trypsin (Promega, Madison, WI, USA) and LysC (Wako, Richmond, VA, USA) at 1:50 and 1:75 enzyme:substrate ratios, respectively. The digestion was quenched with 10% formic acid and the resulting peptides were cleaned-up in an automated fashion using the AssayMap Bravo platform (Agilent Technologies) with corresponding AssayMap C18 reverse-phase column. The eluate was dried and resolubilized in 1% FA to achieve a concentration of 1µg/µL.

### Serum proteome profiling

All spectra were acquired on an Orbitrap HFX mass spectrometer (Thermo Fisher Scientific) operated in data-independent mode (DIA) coupled to an Ultimate3000 liquid chromatography system (Thermo Fisher Scientific) and separated on a 50 cm reversed phase column packed in-house (Agilent Poroshell EC-C18, 2.7um, 50cmx75um). Proteome samples were eluted over a linear gradient of a dual-buffer setup with buffer A (0.1%FA) and buffer B (80%ACN, 0.1%FA) ranging from 9-40% B over 95 min, 40-55% B for 4 min, 55-95% B in 1 min, and maintained at 95% B for the final 5 min with a flow rate of 300 nl/min. DIA runs consisted of a MS1 scan at 60 000 resolution at m/z 200 followed by 30 sequential quadrupole isolation windows of 20 m/z for HCD MS/MS with detection of fragment ions in the OT at 30 000 resolution at m/z 200. The m/z range covered was 400–1000 and the Automatic Gain Control (AGC) was set to 1e6 for MS and 2e5 for MS/MS. The injection time was set to 100ms for MS and ‘auto’ for MS/MS scans.

### Data analysis

Spectra were extracted from the DIA data using DIA-NN (version 1.7.12) without a spectral library and with “Deep learning” option enabled. The enzyme for digestion was set to trypsin and one missed cleavage was tolerated. Cysteine Carbamidomethylation and Methionine oxidation were allowed. The precursor FDR (false discovery rate) threshold was set to 1%. Protein grouping was done by protein names and cross-run normalization was RT-dependent. All other settings were kept at the default values. The gene-centric report from DIA-NN was used for downstream analysis, and quantification was based on unique peptides. When injection replicates were available, the median of these values was used. All downstream analyses were carried out in R [13]. For all proteins the concentration was estimated based on a set of reference proteins using a log-log model. Significant proteins were determined using results from a two-sided Student’s t-test, with a p-value cutoff of 5e-2. Unsupervised hierarchical clustering was performed using Ward’s algorithm with euclidean clustering distance.

## Results

For our proteomics analysis we analyzed 33 patients, selected to obtain two comparable groups in terms of sex (73.3% males), pharmacological treatment and comorbidities, as well as age as much as was possible (median age 71 ±7.6 vs 65 ±9.8, survivor vs deceased) (Suppl. Tables 1, 2). Out of the 17 survivors, 14 had blood withdrawn at 3 timepoints, and 3 patients at only 2 timepoints. In the non-survivor group of 16 patients, 6 patients had blood sampled at 3 timepoints, whereas 5 had 2 timepoints, and 5 had only 1 timepoint (Fig. 1). Following a serum proteomics sample preparation workflow optimized in our group and by others [14, 15] we set out to process all 81 samples simultaneously to avoid introducing batch effects which may confound the results. Data-independent acquisition was performed by analyzing all samples in a randomized order to also further avoid batch effects. The used cohort and the experimental approach are schematically summarized in Figure 1 and further described in the materials and methods section.

**Figure 1.**
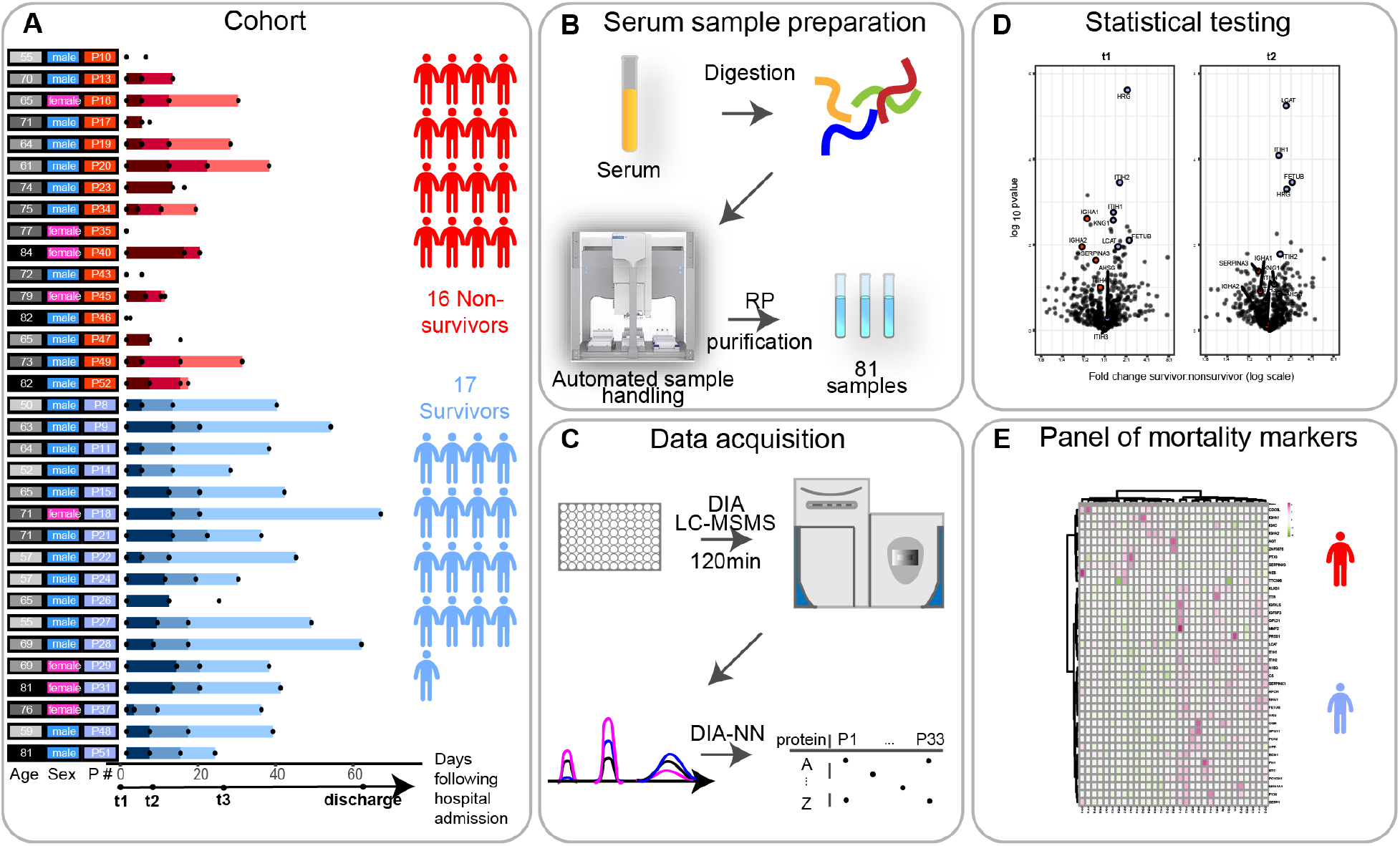
Scheme of the cohort and timing of the blood sample collection, based on each patient’s admission to the hospitalization. A. Serum samples were collected from 33 individuals (17 survivors, 16 deceased) diagnosed with SARS-CoV-2 infection, at one, two or three timepoints following their admission to the clinic (t0). The timepoints t1, t2 and t3 represent blood collections at 96h, 14 days and later than 14 days after they arrived at the ICU of the hospital. The date of discharge from the hospital is recorded here, although no blood was collected at that moment. The numbers represent the elapsed number of days starting from t1 for each patient. These are represented by color gradients ranging from dark to light the longer the duration of the stay in the clinic. Patients where no color timeline is represented, indicate cases for which no consecutive temporal collection points were available. Patient descriptors including age and gender as well as indexes used throughout this report are provided to the right of the timelines and color coded. Patient’s age is binned (10y/bin) and the darker the greyscale in column 1 the older the patient. The gender of each patient is marked in column 2 as blue and pink. Patient indexes are color-coded for patient outcomes, with survivors in blue and deceased SARS-CoV-2 patients in red in column 3. **B**. Serum samples were proteolytically digested and the resulting tryptic peptides were purified using reverse-phase cartridges on an autosampler robot. **C**. The samples were analyzed by LC-MS/MS applying a data-independent acquisition strategy. Spectra were extracted using DIA-NN yielding as a measure abundance for each protein. **D**. Two-sided Student’s t-tests were performed in order to determine significantly regulated proteins comparing survivors and non-survivors. **E**. These differentially regulated proteins were found to be largely functionally related, and define a potential panel of mortality markers, by which we can stratify patients that might ultimately overcome or succumb from the infection, which can be diagnosed already at an early timepoint in the clinic.

In total, and after removal of the numerous detected variable immunoglobulin protein fragments (Suppl. Table 3), we could quantify a mean number of 452 proteins per sample (min=302, max=578) (Fig. 1. B, C). In serum proteomics it has been well established that the total intensity of a protein in label free quantification (i.e. LFQ- or IBAQ-values) can be used as a proxy for protein concentrations. To better relate the abundance of serum proteins to clinical data, we converted the median log label-free quantified values per protein from our mass spectrometry experiments into serum protein concentrations. For this conversion, we performed a linear regression with 22 known reported average values of proteins in serum (A2M, B2M, C1R, C2, C6, C9, CFP, CP, F10, F12, F2, F7, F8, F9, HP, KLKB1, MB, MBL2, SERPINA1, TFRC, TTR, VWF) [16]. This analysis yielded a sensible correlation coefficient of R^2^=0.78 between the proteomics concentration measurements and the average values reported in literature (Suppl. Fig. 1). We therefore decided to convert all mass spectrometric values by using this concentration scale [mg/dL] (Suppl. Table 5).

A first look at the proteins present in our serum samples across all patients revealed a few potential clear outliers. At single timepoints and in single patients several proteins originating from either red blood cells (e.g. HBA1, HBB, CA1, CA2, PRDX2) or fibrinogen (e.g. FGA, FGB and FGG) were extremely prominent (Suppl. Fig. 2). These features are more often observed in serum proteomics and are likely caused by sample preparation artefacts [17]. Fortunately, they do not negatively affect the abundance measurements of the other serum proteins. Therefore, we decided to exclude a panel of well-described RBC contaminants (Suppl. Table 4) from all further analyses. We then sought to determine the differences in serum proteome expression stratifying survivors and deceased patients. For this, at each timepoint separately, we performed a two-sample Student’s t-test and identified proteins significantly differentially expressed between survivors and non-survivors (Fig. 2 A; Suppl. Table 6). At t1, 42 proteins were found to be significantly differentially expressed (28 higher in survivors, 14 higher in non-survivors) taking as threshold a p-value of 0.05. At t2, 30 proteins were found to be differentially expressed (19 higher in survivors, 11 higher in non-survivors). Finally, at t3, 19 proteins were significantly different (10 higher in survivors, 9 higher in non-survivors). In our dataset, 2 proteins were significantly different between survivors and non-survivors at all timepoints (HRG and HPR). Of note, at the latest timepoint, t3, we unfortunately could include only a low number of non-survivors. To illustrate this, the overlap in proteins significantly different between survivors and non-survivors at the first two timepoints was 9 (HRG (p-value at t1: 2.42e-6; t2: 5e-4; t3: 3.27e-2), FETUB (p-value at t1: 7.86e-3; t2: 3.49e-4; t3: 2.51e-1), ITIH1 (p-value at t1: 1.75e-3; t2: 8.18e-5; t3: 1.16e-1), ITIH2 (p-value at t1: 3.52e-4; t2: 1.65e-2; t3: 2.83e-1), HPR (p-value at t1: 2.17e-2; t2: 3.66e-2; t3: 3.25e-3), SERPINA3 (p-value at t1: 2.31e-2; t2: 4.11e-2; t3: 2.67e-1), LCAT (p-value at t1: 1.11e-2; t2: 5.64e-6; t3: 7.98e-2), IGFALS (p-value at t1: 1.16e-2; t2: 8.78e-3; t3: 4.79e-1), IGFBP3 (p-value at t1: 3.76e-3; t2: 1.37e-2; t3:6.74e-1)). The nature of the first two timepoints as well as the number of patient samples available at these timepoints however led us to focus first on these as they would enable the most appropriate comparison (at t1 there are 16 survivors and 15 non survivors, compared to t2 with 15 *vs* 11 and t3 with 16 *vs* 6). We did choose to first focus on the data for t1, as this represents also the most narrowly defined timeframe compared to t2 and t3. Although we thus do deliberately not focus on timepoint t3, due to the lower statistics, we did observe at this last timepoint that several neutrophil originating proteins, such as MPO, PRTN3 and LCN2, indicated in yellow in Fig. 2A, were more abundant in the non-survivors. Four proteins that were clearly significant at timepoint 1 but that did not pass the significance threshold at timepoint 2, and are of particular interest were FN1 (p-value at t1: 4.82e-3; t2: 1.61e-1; t3: 4.37e-1), IGHA1 (p-value at t1: 2.45e-3; t2: 1.21e-1; t3: 2.67e-1), IGHA2 (p-value at t1: 1.11e-2; t2: 7.9e-1; t3: 1.25e-1), and KNG1 (p-value at t1: 2.67e-3; t2: 8.29e-2; t3: 5.68e-1).

**Figure 2.**
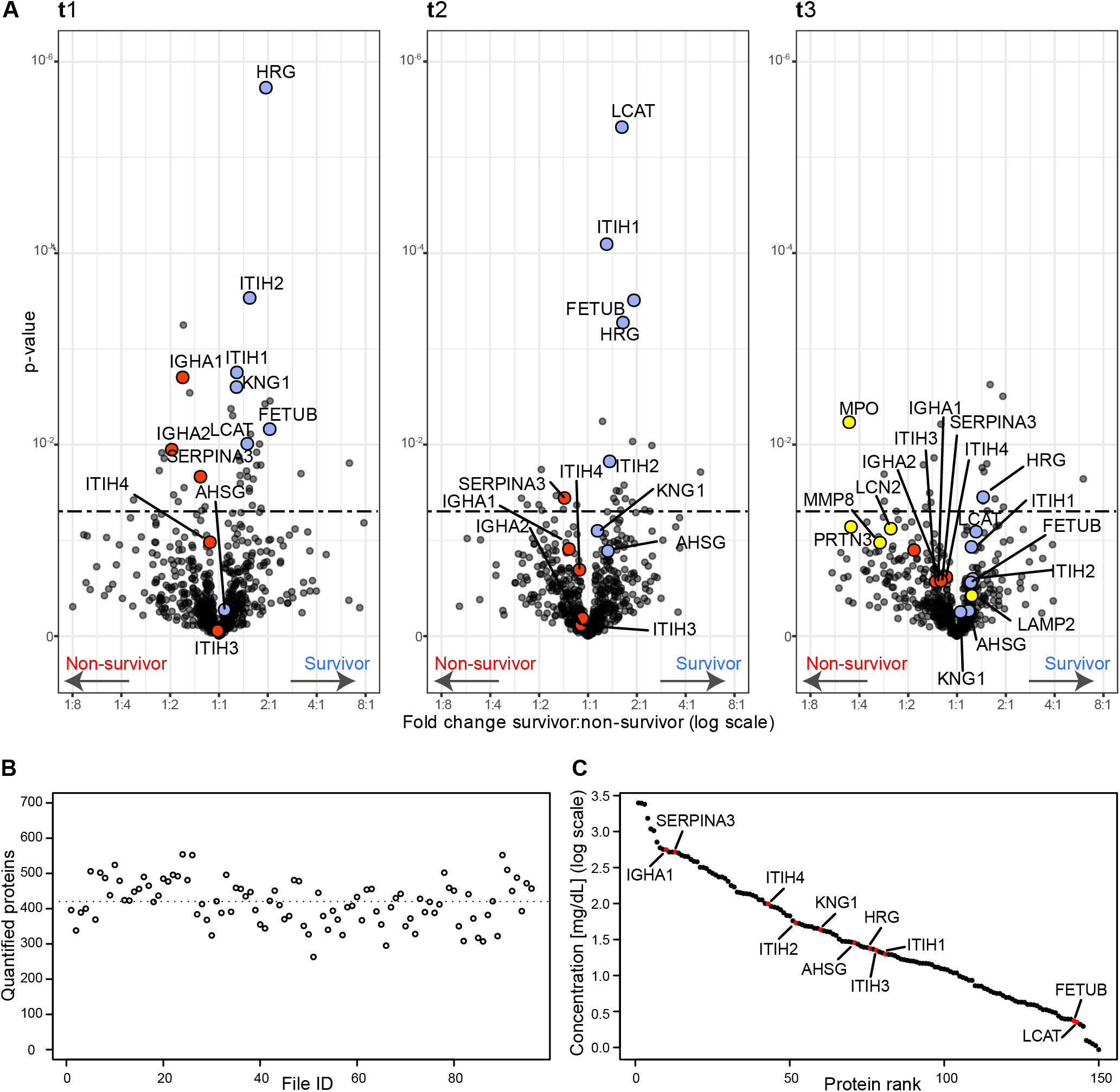
Serum proteins that are differentially abundant in survivors and non-survivors per timepoint. **A** Volcano plots showing the fold change and associated p-values. For each timepoint t1, t2 and t3, a two-sided Student’s t-test was performed to identify the significance of the differentially abundant serum proteins (significance threshold at p-value 0.05 indicated as dashed line). Proteins discussed here are represented in red if higher in deceased patients and in blue if showing an increase in surviving patients. In the Volcano plot of t3 some proteins of neutrophil origin are highlighted in yellow. **B**. The number of quantified proteins per sample, with the dotted line indicating the mean (452 proteins quantified on average). **C**. The quantitative MS-based data converted to the concentration scale for the 150 abundant serum proteins that are identified and quantified in all samples. Proteins we discuss as potentially stratifying survivors and non-survivors are marked in red (AHSG, FETUB, KNG1, HRG, ITIH1, ITIH2, LCAT, SERPINA3, IGHA1, IGHA2, ITIH3, ITIH4) and thus span the entire covered dynamic range.

Thus, assuming our data represents reasonably accurate protein concentrations, we next looked at the potential functional relationships between the differentially abundant proteins from our comparative analysis.

With a focus on the proteins showing differences in abundance at timepoint 1, we applied an unsupervised clustering approach and found as expected that the differentially regulated proteins that resulted from the analysis of timepoint 1 samples show a clear cluster that is distinct between the survivors and some non-survivors (Fig. 3 A). These same differentially regulated proteins also perform relatively well at timepoint 2 to stratify between patient outcomes (Fig. 3 B).

**Figure 3.**
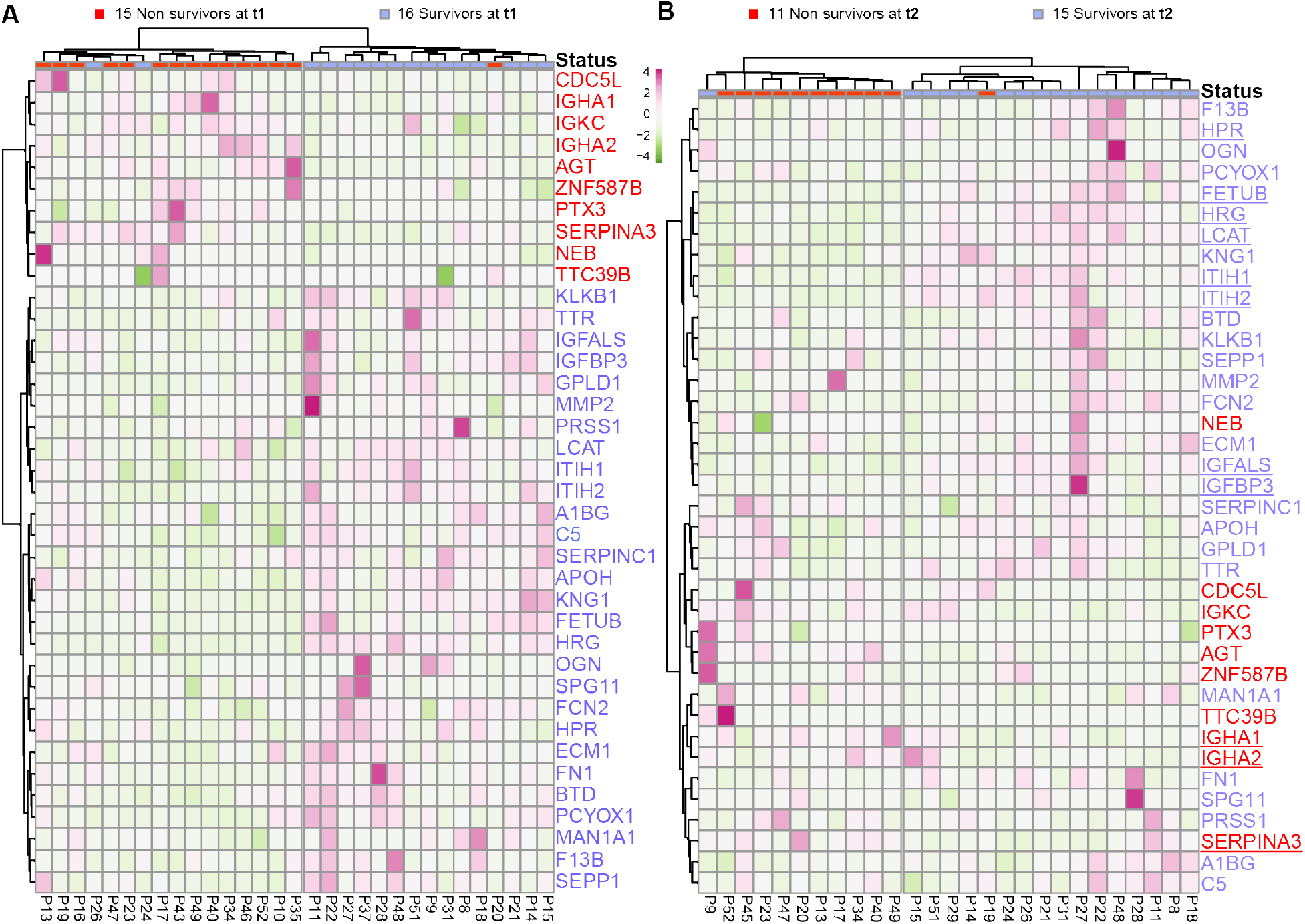
Surviving and deceased SARS-CoV-2 patients can be distinguished by a small panel of abundant serum proteins, already at t1. **A**. Proteins identified as differentially abundant at timepoint 1 are shown here to completely cluster samples with respect to patient outcome (16 survivors VS 15 non-survivors at timepoint 1). Proteins are annotated by their gene names, and those indicated in blue show higher concentrations in survivors whereas proteins in red show higher concentration in non-survivors. **B**. The differentially abundant proteins at timepoint 1 (as shown in A) are used to distinguish the 15 survivors from 11 non-survivors at timepoint 2. Underlined proteins in B designate the proteins that were found to be regulated at both timepoints. Although the cohort consisted of 17 survivors and 16 non-survivors, at each timepoint we missed for a few patients a blood sampling point, which then also could not be included in the clustering and is indicated at the top of the dendrograms.

## Discussion

In this study we prospectively performed serum proteomics in moderate-to-severe COVID19 patients admitted to Respiratory and Intensive Care Units because of respiratory failure. Our serum proteomics data provided quantitative information about the abundance of about 300-400 serum proteins on average per patient and per timepoint, and thus provided quantitative information about changes in protein abundances over disease progression per patient, but also information about serum proteins being more or less abundant when comparing the group of survivors and those that died.

Analyzing the quantitative serum proteomics data, we were intrigued by the fact that about two dozen of the highly abundant proteins seemed to be significantly more (or less) abundant in the serum of survivors *versus* the group of deceased patients. Some of the proteins being more abundant in the survivors contained the structurally and functionally related type 3 cystatins fetuin-B (FETUB), the histidine-rich glycoprotein (HRG) and kininogen (KNG1), the inter-*alpha*-trypsin inhibitor isoforms (ITIH1) and (ITIH2) and phosphatidylcholine-sterol acyltransferase (LCAT). The proteins being more abundant in the deceased patients contained alpha-1-antichymotrypsin (SERPINA3), the immunoglobulins IgA (IgA1 and IgA2), the inter-*alpha*-trypsin inhibitor isoforms ITIH3 and ITIH4. Moreover, fibronectin (FN1) showed a higher abundance in non-survivors, while decreasing in survivors. The differences for this panel of proteins were consistently observed at several of the sampling timepoints and display consistent trends over time and may therefore potentially be considered as a panel that can be used for mortality risk assessment. Before discussing whether this panel may be sufficiently predictive, we first discuss these findings by discussing the functional role and relationship of these serum proteins, all of which are amongst the most abundant.

FETUB, HRG and KNG1 are in our data some of the most distinctive differentially abundant serum proteins between survivors and non-survivors, with their levels all about a factor 2-4 more abundant in the survivors, at almost all 3 timepoints sampled. The fourth family member is fetuin-A (AHSG), which although not significant in our testing, followed a similar trend (see Figure 4A). FETUB, HRG, KNG1 and AHSG are all structurally and functionally related type 3 cystatins. They are all four closely located to each other on the human chromosome 3 (Figure 4E). They share some sequence homology, all containing either 2 or 3 alike cystatin domains, and KNG1 and HRG share also a His-rich domain. HRG is present in human plasma at approximately 75-150 mg/mL in healthy donors and has been implicated in quite a variety of biological functions [18]. HRG was also found to be a negative acute phase reactant and circulating HRG levels were found to be significantly lower during acute inflammation and in patients with systemic lupus erythematosus. It has been suggested that HRG may play a critical role in recognizing common molecular “danger” signals in the innate response that protects against tissue damage and pathogen invasion as well as aiding wound healing. Still, a clear function of HRG in plasma has not been defined, instead it has been termed an important multifunctional protein, or even swiss-army knife, due to its ability to interact with a wide range of small molecules and other plasma proteins as reviewed by Poon et al. Fetuin-A, referred to as alpha-2-HS-glycoprotein (AHSG) is also an abundant and important plasma protein, albeit also defined as a multifunctional protein [19, 20]. FETUB is a close paralog of HRG; alignment of these two genes reveals 35% identity. Also, its functional role in plasma is still to some extent obscure and certainly also multifunctional. Finally, Kininogen-1 (KNG1), also known as alpha-2-thiol proteinase inhibitor, is maybe best known as the precursor for the low molecular weight peptide bradykinin. It contains 3 cystatin domains and shares a histidine-rich domain with HRG. Thus, AHSG, FETUB, HRG and KNG1 share domain structure (cystatin-domains), chromosome localization and functionality. In our serum proteomics data, the abundances of AHSG, FETUB, HRG and KNG1 are found to be highly correlated in each of the sampled COVID-19 patients and are consistently higher in the survivors. In general, it does however seem that the abundance levels of these type 3 cystatins remain fairly constant during disease progression.

**Figure 4.**
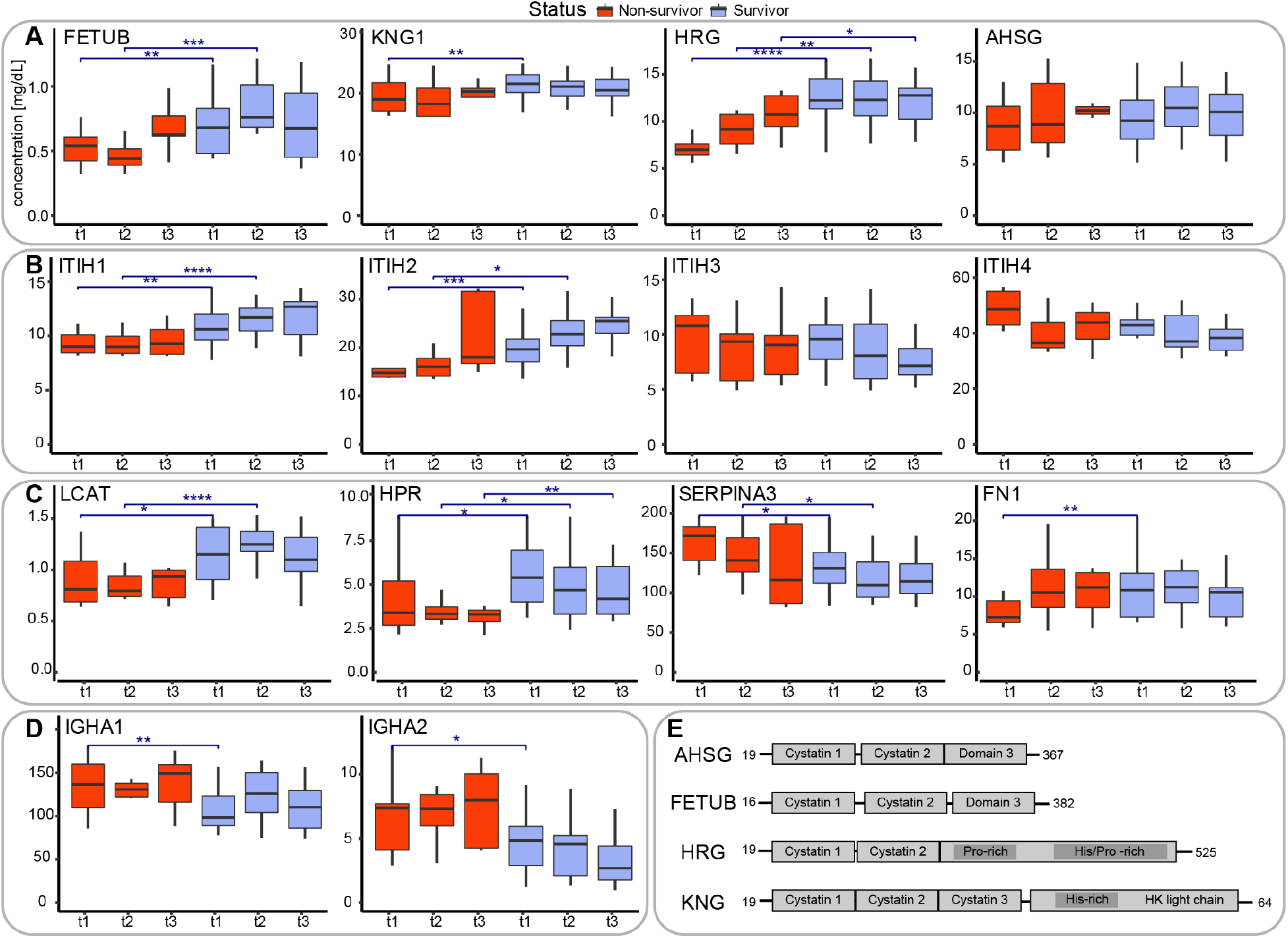
Serum abundance and structural and functional description of proteins being differentially abundant in survivors vs. non-survivors. **A**. Serum abundance of the four type-3 cystatins, comparing survivors (blue) with non-survivors (red). At each timepoint the abundance of these cystatins is higher in the survivors compared to the non-survivors. Comparisons between survivors and non-survivors at respective timepoints are denoted as significant using asterisks: * p <= 0.05, ** p <= 0.01, *** p <= 0.001 and **** p <= 0.0001. **B**. Serum abundance of the four abundant inter-α-trypsin inhibitors (IαI), with clear opposing trends between ITIH1 and ITIH2 as well as between ITIH3 and ITIH4. At each timepoint the abundance of ITIH1 and ITIH2 is higher in the survivors compared to the non-survivors, whereas for ITIH3 and ITIH4 the opposite holds. **C**. Serum abundance of other putative mortality indicators: phosphatidylcholine-sterol acyltransferase (LCAT), haptoglobin-related protein (HPR), alpha-1-antichymotrypsin (SERPINA3), and fibronectin (FN1). **D**. Profiles of the IgA immunoglobulin variants IgA1, IgA2, both less abundant in survivors. **E**. Schematic domain-structures of the type-3 cystatins showing their sequence homology. Cystatin domains as well as His/Gly and His/Pro domains are depicted as boxes.

Of interest, two recent studies have hypothesized that these abundant plasma proteins may indeed represent mortality markers. Firstly, Hong et al. [9] used a multiplexed antibody-based affinity proteomics assay to screen 156 individuals aged 50–92. This dataset revealed a consistent age association for the histidine-rich glycoprotein (HRG). They concluded, by validating this finding in several additional data sets (N = 3,987), that the histidine-rich glycoprotein associates with age and risk of all-cause mortality, whereby a GWAS analysis indicated that particular mutations in HRG may influence the mortality risk. Secondly, Wozniak et al. analyzed a cohort of around 200 patients by serum proteomics and metabolomics to assess whether there are potential mortality risk biomarkers for patients that suffered a *Staphylococcus aureus* bacteremia. The statistically most significant prediction marker they observed was FETUB, which was consistently higher in survivors than in diseased patients. These data and our current data therefore may indicate that high serum levels of the cystatins fetuin-A (AHSG), fetuin-B (FETUB), the histidine-rich glycoprotein (HRG) and kininogen (KNG1) are general positive survival factors, independent of the pathogen that induces the disease.

### The family of Inter-α-trypsin inhibitors (IαI) as putative mortality markers

In our dataset a set of other functionally related proteins stand out, all belonging to the family of inter-α-trypsin inhibitors. Four of these are abundantly present in our serum dataset, namely ITIH1, ITIH2, ITIH3 and ITH4. Notably, ITIH1 and ITIH2 are statistically significant differently abundant in between survivors and non-survivors, but all four inhibitors also show opposite trends during disease progression (Fig. 4 B). Notably, the abundance levels of ITIH1 and ITIH2 correlate extremely well in each serum sample analyzed, which we actually expected as they are known to form in serum an approximately 225-kDa complex, named IαI, containing Bikunin next to ITIH1 and ITIH2. Bikunin is a proteoglycan with a chondroitin sulfate (CS) chain attached to the protein core of approximately 20 kDa, also termed AMBP. We observe a strong positive correlation between ITIH1 and ITIH2, providing confidence in our quantification. ITIH1 and ITIH2 are consistently higher in survivors than in non-survivors. Additionally, the levels of these two proteins increase during disease progression, both in the survivors and non-survivors. In sharp contrast, the abundance levels of ITIH3 and ITIH4 (that do not form a complex) show a decreasing trend during disease progression, and moreover these proteins are, although just below our statistical cut-off, consistently lower in abundance in survivors than in non-survivors. In a comprehensive recent review by Lord et al. [21] the structural organization and functional role of the members of these inter-α-trypsin inhibitors (IαI) is described. The exact role of these proteins in serum is not fully clear, although they all exhibit matrix protective activity through protease inhibitory action. Next, IαI family members interact with the extracellular matrix and most notably hyaluronan, inhibit complement, and provide several cell regulatory functions. Notably, a reduction in circulating IαI has been reported in patients with sepsis which correlated with increased mortality rates [22]. Moreover, administration of exogenous IαI has been shown to lead to reduced mortality suggesting a protective role of specific IαI family members [10]. It is somewhat difficult to relate these earlier findings with our data, as in these studies, using a broad-spectrum antibody, no distinction was made between the different inter-α-trypsin inhibitor family members. Still, as in our samples ITIH4 is by far the most abundant in serum, it is nice to see that its abundance is indeed lower in the non-survivors, consistent with the findings of Lim et al. and Opal et al., where inter-alpha inhibitor proteins were found to be reduced in severe sepsis, and failure of recovery of IαIp levels over the course of sepsis was associated with mortality. Our data confirm that hypothesis when considering ITIH4 and ITIH3, but notably the opposite is observed for ITIH2 and ITIH1. In summary, the four related inter-α-trypsin inhibitor members possibly provide a panel for monitoring disease outcome in a range of pathogen caused diseases, amongst them COVID-19. Of note, in the context of sepsis IαI improves endothelial inflammation while their levels are inversely associated with the levels of endothelial dysfunction biomarkers VCAM-1 and ICAM-1 [23]. Endothelial dysfunction is a feature of COVID-19 [24] and we have previously observed that high levels of ICAM-1 [25] and VCAM-1 [26] at admission are associated with the mortality of our COVID-19 patients. Our new data suggests that IαI may provide protection from endothelial complications of COVID-19, thereby potentially improving survival.

Finally, IαI has been used in the treatment of inflammatory conditions such as sepsis (one of the most common causes of death for COVID-19 patients [27]) but also for Kawasaki disease, which has recently been associated with SARS-Cov-2 infection in children [28].

### Other putative mortality predictors

Next to the family of type-3 cystatins and the family of inter-α-trypsin inhibitors, only a few more serum proteins did stand out significantly as potential mortality predictors. These included the phosphatidylcholine-sterol acyltransferase (LCAT), an enzyme involved in the extracellular metabolism of plasma lipoproteins, which showed a trend similar to ITIH1 and ITIH2, consistently higher in survivors than in non-survivors and increasing in abundance during disease progression, both in the survivors and non-survivors. Reversely, the family of immunoglobulin A, IgA1 and IgA2, seemed to be more abundant in non-survivors as well as alpha-1-antichymotrypsin (SERPINA3) (Fig. 4 C, D). Thereby angiotensin and alpha-1-antichymotrypsin showed a decrease in abundance over time, while the IgA levels seemed to remain more constant over time.

### Limitations of the study and comparison with related recent COVID-19 plasma proteome studies

Although the panel of plasma protein markers we here define as putative mortality indicators is very significant in stratifying the survivors from the non-survivors affected by COVID-19, our study has a few limitations. First of all, it is still a rather small cohort of patients. Moreover, patients’ characteristics mainly related to medical history and treatments can affect the measured outcomes. However, we also believe that the prospective longitudinal design adopted in the study and the rigorous clinical follow up increase the strength of the results, particularly in relation to clinical outcomes.

With this in mind, our study should ideally be compared to alike studies on other independent cohorts. In the last months, the research efforts on COVID-19 have expanded enormously. In this period, two related plasma proteomics studies have appeared studying COVID-19 patients, generating data comparable to ours, but with different research questions and thus also different study design. Still, the outcome of these studies and their conclusions can be compared with our data.

First of all, Demichev *et al*. in a study coordinated by the PA-COVID-19 study group [3], measured the plasma proteome of 139 hospitalized patients, and followed the time-dependent progression of COVID-19 through different stages of the disease, combining several diagnostic clinical parameters and plasma proteomes. They used the clinical parameters to classify the patients in cohorts of increasing severity and followed changes in the patients’ clinical parameters and plasma proteomes over time. From their data they were able to define signatures for disease states as well as observe age-related plasma proteome changes. Relevant to our work, they report that low plasma levels of 54 proteins could be associated with disease severity. Comparing that list with the list of 28 proteins that are found to be significantly lower in non-survivors *versus* survivors at t1 in our work, we observe a large overlap, including HRG, FETUB, AHSG, IGFALS, GPLD1, LCAT, TTR, SERPINC1, HPR, ITIH1 and ITIH2, all lower in abundance in the plasma of severe COVID-19 patients. The list of 54 proteins of Demichev *et al*. is larger than ours, but their dataset also includes proteins we disregarded as RBC contaminants, such as variants of hemoglobin (HBD, HBB, HBA1) and carbonic anhydrase, or excluded from our analysis for other reasons, such as albumin. Reversely, we also examined the list of proteins they state as being of high abundance in the plasma of patients with poor prognosis and also observe a substantial overlap. As was the case for low abundant proteins in patients with poor prognosis, their list includes proteins we disregarded, such as the fibrinogens FGA, FGB and FGG, as these levels may be affected by the sampling. Other proteins they note are also in our list of proteins being higher in non-survivors at t1, notably SERPINA3, AGT, ITIH3, and ITIH4.

From their clinical and plasma proteome data they ultimately defined a very narrow panel of proteins predicting future worsening of COVID-19 disease. The four proteins defined by them to be indicative of poor prognosis when their plasma levels are low were AHSG, HRG, ITIH2 and PLG. Pleasingly, except for PLG all these proteins were also found by us to be substantially lower in the non-survivors than non-survivors. Surprisingly, ITIH1 was not mentioned by Demichev *et al*. in this panel, although the protein levels of this protein are known to directly correlate with that of ITIH2. Conversely, they defined 7 proteins whereby high abundance in plasma would be indicative of poor prognosis, including SERPINA3 and AGT. Both these latter proteins are also part of the small putative mortality panel we define here and are indeed lower in abundance in our non-survivors. The data of Demichev *et al*. are therefore in excellent agreement with our findings.

Another recent plasma proteome profiling study related to ours is from Geyer et al [29]. They primarily studied differences between the plasma proteomes of 31 COVID-19 patients *versus* 263 PCR-negative controls. In that analysis Geyer et al. found that the protease inhibitors SERPINA3, ITIH3 and ITIH4 were increased in plasma abundance in COVID-19 patients, when compared to controls, whereas the histidine-rich glycoprotein (HRG) and fibronectin (FN1) were decreased. In our study we did not compare the COVID-19 plasma proteomes to PCR-negative controls but focused on ICU-hospitalized survivors *versus* non-survivors. Still, we also find that in the survivors SERPINA3, ITIH3 and ITIH4 were lower in abundance and HRG and FN1 higher in abundance, when compared to the non-survivors, confirming that low levels of e.g. HRG are potentially a hallmark of COVID-19 disease diagnosis and disease severity.

Next, Geyer et al. also followed the protein abundances in the 31 COVID-19 patients over time. They measured longitudinal trajectories of 116 proteins (a list including many immunoglobulin variants, that we however chose to exclude from our analysis) that significantly changed over a disease course of up to 37 days. Their more in-depth study, albeit with a similar size cohort, but with more frequent sampling at a greater number of timepoints, can be directly compared to the data for our 33 patients, sampled at just three timepoints. It is striking and pleasing to see that the trends observed are in very good agreement, certainly for the small panel we define here as putative mortality markers. Pleasingly the trends observed for all the four abundant inter-α-trypsin inhibitors (ITIH1, ITIH2, ITIH3 and ITIH4) are alike in both studies, with an increase over time for the first two and a decline in the latter two. Moreover, they observe the plasma proteins HRG, FETUB, KNG1, LCAT, AHSG and FN1 to increase over time in abundance, thus suggesting their regulation during COVID-19 disease development. Finally, SERPINA3, found to decrease over time in our study was observed to decrease over time by Geyer et al. as well.

Thus, although the study designs as well as the patient cohorts were completely different, our dataset compares extremely well with that reported independently by Demichev *et al*. and Geyer *et al*., with a similar small panel of serum proteins presenting hallmarks for disease diagnosis, development and severity. The huge consistency in the findings provides credibility to these independently made observations, especially considering the still modest number of patients all three studies tackled.

In summary, in our study comparing one-by-one the serum proteomes of a closely matched group of survivor and non-survivor COVID-19 patients admitted to the ICU, we were able to extract a small subset of proteins that can be used to predict the disease outcome, already at an early stage, i.e. shortly after admission to the ICU. This set of mortality indicators consists largely of functionally related proteins, namely several type-3 cystatins and the family of inter-α-trypsin inhibitors. Although our patient cohort was rather small, our observations and conclusions are in pleasing agreement with two other, independent, recent serum proteome studies, which basically define the same set of proteins as either markers or disease severity or mortality. Finally, we hypothesize, based on comparison with existing literature, that the here defined mortality risk panel of proteins may predict mortality not specifically for COVID-19 patients, but also in other pathogen caused diseases, and even more in general also in aging.

## Supporting information

checklist

## Data Availability

The mass spectrometric proteomics data have been deposited at the ProteomeXchange Consortium via the PRIDE partner repository with the dataset identifier PXD024707.

http://www.ebi.ac.uk/pride

## Data Availability

The mass spectrometric proteomics data have been deposited at the ProteomeXchange Consortium via the PRIDE partner repository [30] with the dataset identifier PXD024707.

## Author Contribution

FV and AJRH conceptually conceived and designed the analysis performed in this proteomics study, analyzed the serum proteomics data and wrote the paper. GC, MC and SS conceived and designed the original clinical studies. GC, MC, SS, CAV, AP, and FVDS assisted in data interpretation and provided critical revision of the manuscript providing key intellectual content. OZ, FVDS and PR provided clinical data and contributed to the study design and critically revised the manuscript. FF and LM conducted the molecular analyses to exclude the presence of the virus in the analyzed sera. FV executed all proteomics experiments. HvdT provided bioinformatics support and assisted in the data analysis. AJRH supervised the proteomics analysis performed in this study.

## Acknowledgements

We acknowledge support from the Dutch Research Council (NWO) funding the Netherlands Proteomics Centre through the X-omics Road Map program (project 184.034.019) and the EU Horizon 2020 program INFRAIA project Epic-XS (Project 823839).

## Supplementary Figures

**Suppl. Fig. 1.**
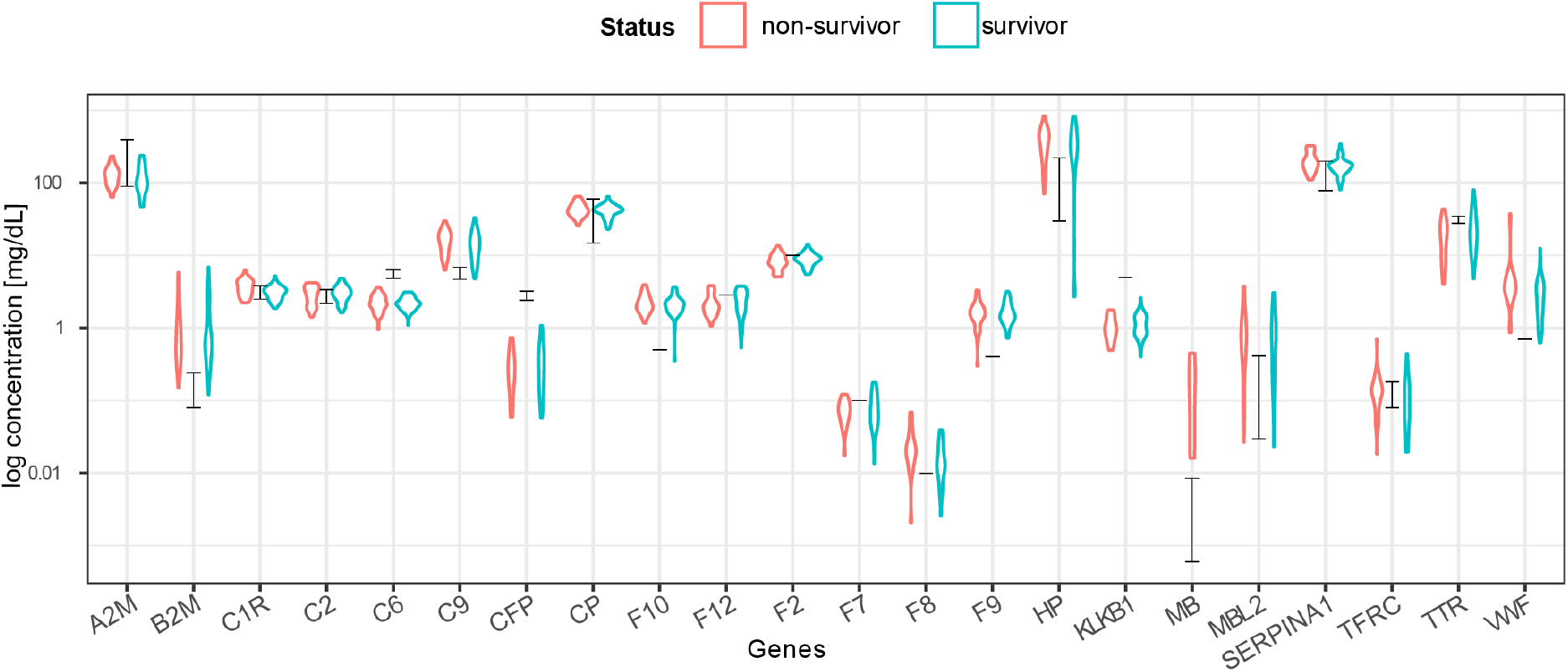
Concentration estimates based on MS-based label free quantification correlate well with reported plasma concentration data. MS label-free quantified intensities were converted to a clinical concentration scale (expressed in mg/dL) based on measurements of 22 reference proteins [16] The black bars indicate the ranges from the reference concentrations. Individual protein measurements correlate well with the reference concentrations, whereas the red and blue violin plots indicate the estimated concentrations for the non-survivor and survivor groups.

**Suppl. Fig. 2.**
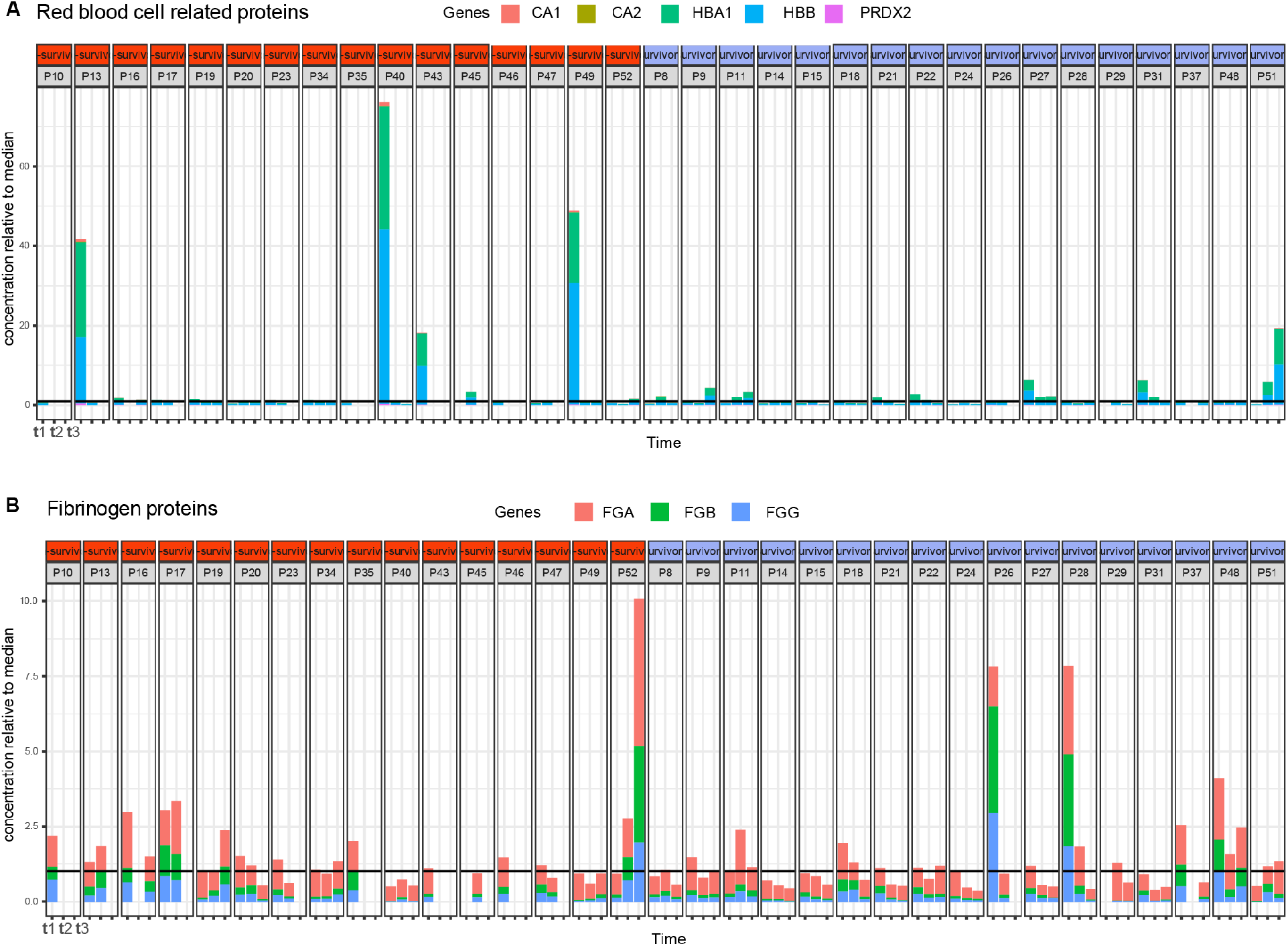
Abundance of a selection of **A**. red blood cell related contaminants displaying high variability and outliers, mainly at timepoint 1 of patients P13, P40, P49. **B**. Fibrinogen-related contaminants. The black horizontal line indicates the median concentration, which has been normalized to the value ‘1’, so for instance at timepoint t1, patient P13 red blood cell protein concentration is about 40 times what is expected in plasma.

**Suppl. Fig. 3.**
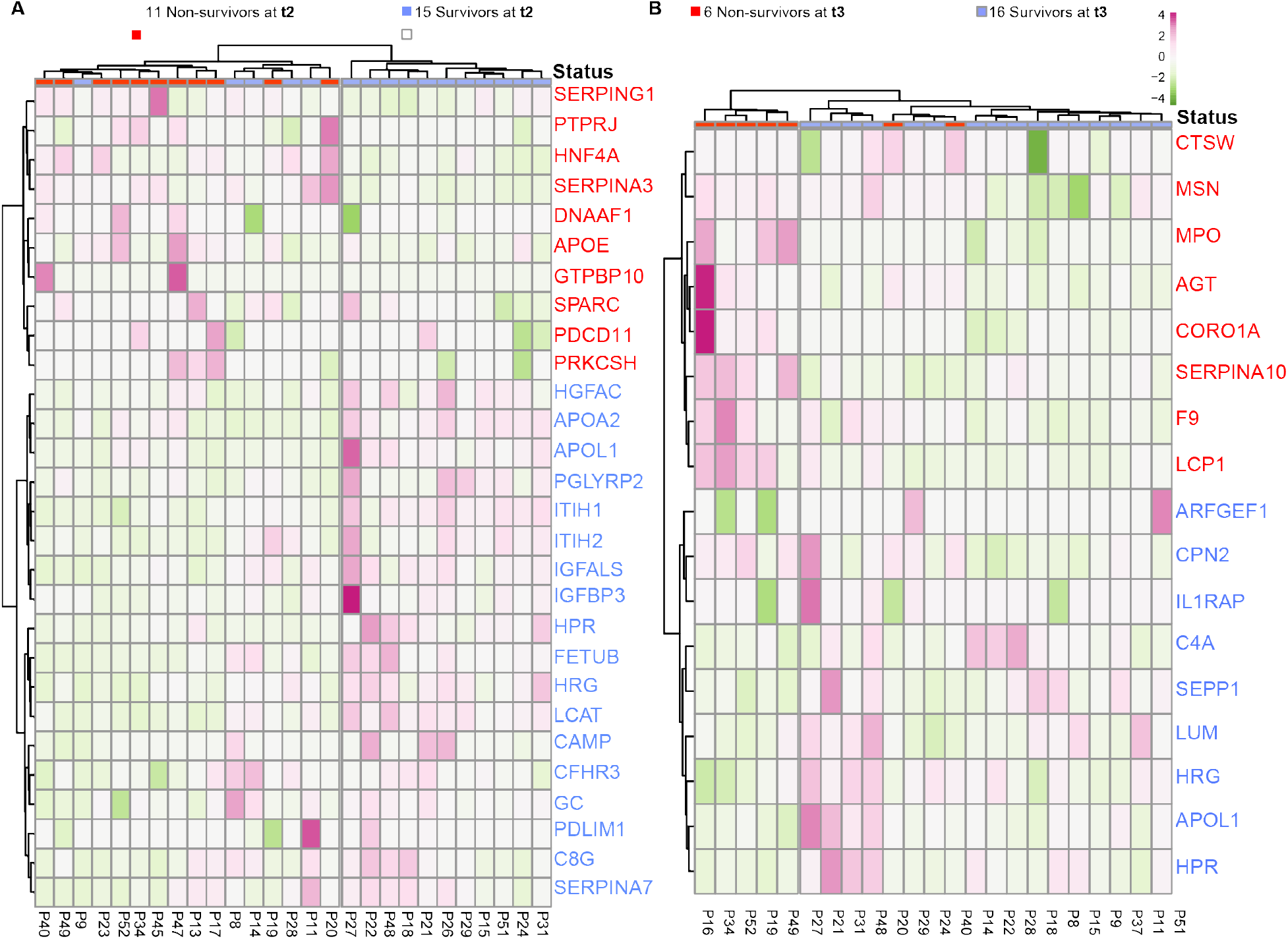
Surviving and deceased SARS-CoV-2 patients can be reasonably well distinguished by a small panel of statistically significant serum proteins, at t2 and t3. **A**. Proteins identified as differentially abundant at timepoint are shown here to cluster samples with respect to patient outcome (15 survivors VS 11 non-survivors at t2). Proteins are annotated by their gene names, and those indicated in blue show higher concentrations in survivors whereas proteins in red show higher concentration in non-survivors. **B**. The differentially abundant proteins at timepoint 3 cluster the 16 survivors from 11 non-survivors at t3. Although the full cohort consisted of 17 survivors and 16 non-survivors, at each timepoint we missed for a few patients a blood sampling point, which then also could not be included in the clustering and is indicated at the top of the dendrograms.

## Supplementary Tables

**Suppl. Table 1** Clinical metadata.

**Suppl. Table 2** Baseline characteristics.

**Suppl. Table 3** List of variable immunoglobulins removed from the analysis.

**Suppl. Table 4** List of contaminants removed prior to statistical testing.

**Suppl. Table 5** Protein abundance expressed in clinical concentration range, per patient, per timepoint.

**Suppl. Table 6** Proteins submitted to statistical testing and the results thereof.

